# Overweight in children under five in urban and wealthier households in Bangladesh: trends and inequalities across three Multiple Indicator Cluster Surveys, 2012-13 to 2025

**DOI:** 10.64898/2026.10.22.26366037

**Authors:** Mohammad Khalid Hasan, S. M. Ashraf Ul Hoque, Md Khairul Islam, Ashika Sultana, Tanvir Ahmed, Kamran ul Baset

## Abstract

**Background:** Childhood overweight is often expected to emerge first among urban and wealthier populations during the nutrition transition. However, evidence on temporal trends in childhood overweight and the independent contributions of household wealth and urban residence in Bangladesh remains limited.

**Methods:** We analysed data from 63,972 children aged 0-59 months with valid weight-for-height z-scores (WHZ) from the Bangladesh Multiple Indicator Cluster Surveys (MICS) of 2012-13, 2019 and 2025. Overweight was defined as WHZ >+2 according to the 2006 WHO Child Growth Standards. We estimated survey-weighted prevalence, absolute and relative wealth inequalities, and prevalence ratios for urban versus rural residence using regression models adjusted for household wealth and other measured characteristics. We also assessed anthropometric data quality and conducted sensitivity analyses.

**Results:** National overweight prevalence was 1.6% (95% CI 1.4-1.9) in 2012-13, 2.4% (2.2-2.7) in 2019 and 1.7% (1.5-2.0) in 2025. The 2019 peak was concentrated in urban Dhaka district, where one measurer recorded all 67 overweight cases and WHZ variability was high. Excluding this area, prevalence was 1.5%, 1.9% and 1.7%, respectively. The fully adjusted relative index of inequality for wealth remained similar across surveys: 3.66 (1.93-6.93), 3.55 (2.21-5.71) and 3.35 (2.02-5.54), with no evidence of temporal change (interaction p=0.974). The wealth-adjusted urban-to-rural prevalence ratio was 1.44 (1.01-2.06) in 2012-13 and 1.77 (1.40-2.24) in 2019, then fell to 0.72 (0.54-0.95) in 2025. Mean WHZ decreased by 0.16 (95% CI-0.19 to -0.13) between 2019 and 2025, while wasting prevalence increased from 9.8% to 12.9%. In 2025, 41.4% (35.3-47.7) of overweight children were also stunted.

**Conclusions:** Childhood overweight showed no sustained national increase, but persistent wealth inequalities remained. The changing association with urban residence and concerns about measurement quality highlight the need for continued surveillance and stronger anthropometric quality assurance.

## Introduction

Overweight in early childhood is associated with an increased risk of obesity in adulthood, with potential implications for long-term health [1]. Low and middle-income countries increasingly face a double burden of malnutrition, in which undernutrition and overweight coexist within the same populations [2,3]. This coexistence reflects complex nutritional transitions involving changes in food systems, the availability and consumption of processed foods, and physical activity patterns [3]. The rise in mean body mass index among children and adolescents has accelerated in South and East Asia [4], highlighting the importance of understanding how these broader trends are reflected in younger children. The World Health Assembly set a target of no increase in overweight among children under five years by 2025 [5].

The distribution of childhood overweight may vary according to household socioeconomic position and place of residence. During the nutrition transition, wealthier and urban populations may experience changes in dietary and lifestyle exposures earlier than other groups [2,3]. Evidence from five South Asian countries showed that overweight was more prevalent among children in the richest than in the poorest households [6]. In Bangladesh, analyses of the 2017-2018 Demographic and Health Survey reported overweight prevalence of 2.2%-3.3% among children under five [7,8]. Research among urban school-age children has also documented the coexistence of undernutrition and overweight [9]. However, these findings do not establish whether overweight among younger children in Bangladesh has increased over time or whether observed urban differences persist after accounting for household wealth. Distinguishing these patterns is important because wealth and urban residence may overlap, making their independent associations difficult to determine.

Measurement quality presents an additional challenge when assessing trends in childhood overweight. Overweight is defined by a threshold in the upper tail of the weight-for-height z-score (WHZ) distribution. Random measurement error can increase the dispersion of anthropometric measurements and inflate the proportion of children classified above this threshold [10,11]. Consequently, differences in observed prevalence across surveys may partly reflect variation in anthropometric data quality rather than changes in nutritional status alone [12]. Assessing measurement quality is therefore important when interpreting temporal and socioeconomic patterns in this relatively uncommon outcome.

The Bangladesh Multiple Indicator Cluster Surveys (MICS) of 2012-2013, 2019 and 2025 provide an opportunity to examine childhood overweight across multiple survey rounds using a common analytical framework [13–15]. We aimed to assess trends in overweight among children aged 0-59 months nationally and by household wealth and place of residence. We examined the independent associations of wealth and urban residence, assessed whether these inequalities changed across survey rounds, and characterised the distribution of overweight in 2025, including its coexistence with stunting. We also evaluated anthropometric data quality and conducted sensitivity analyses to assess the robustness of the observed patterns.

## Methods

### Study design and data sources

We conducted a repeated cross-sectional analysis of data from three Bangladesh Multiple Indicator Cluster Surveys (MICS): 2012-13, 2019, and 2025. The surveys were implemented by the Bangladesh Bureau of Statistics (BBS), in collaboration with UNICEF Bangladesh, as part of the global MICS programme [13–16]. The surveys used nationally representative, multistage probability sampling designs to generate estimates of child and maternal health and other population indicators.

Each survey used a two-stage, stratified cluster sampling design. Urban and rural areas within districts constituted the sampling strata, and census enumeration areas served as primary sampling units. Households were selected from sampled enumeration areas. The 2012-13 and 2019 surveys used the 2011 census sampling frame, whereas the 2025 survey used the 2022 census frame and its corresponding urban-rural classification. Fieldwork was conducted from December 2012 to April 2013, January to June 2019, and February to June 2025, respectively.

We included all children aged 0-59 months with a completed child questionnaire and valid weight-for-height z-score (WHZ). The analysis used existing survey data; no additional participants were recruited or measured for this study.

### Study population and anthropometric assessment

All children younger than 5 years in sampled households were eligible for anthropometric assessment under the respective survey protocols. Trained fieldworkers measured children’s weight and length or height using UNICEF-recommended equipment and procedures. Children younger than 24 months were measured in the recumbent position, whereas older children were measured standing. We excluded children without valid anthropometric measurements and those whose WHZ values were flagged as biologically implausible. The participant flow and exclusions by survey round are presented in Supplementary Table S9.

### Outcome definitions

The primary outcome was overweight, defined as WHZ greater than +2 standard deviations relative to the 2006 WHO Child Growth Standards [17]. We used the WHO-standardised anthropometric variables provided in the MICS datasets and excluded observations flagged as implausible in accordance with WHO and UNICEF recommendations [18].

Stunting was defined as height-for-age z-score (HAZ) below -2 standard deviations. Concurrent overweight and stunting were defined as WHZ greater than +2 and HAZ below -2 in the same child. Overweight without stunting was defined as WHZ greater than +2 and HAZ of -2 or higher among children with valid values for both indicators.

Mean WHZ was also analysed as a continuous outcome to assess temporal changes across the weight-for-height distribution.

### Exposures and covariates

The principal exposures were household wealth and place of residence. Household wealth was measured using the MICS wealth index, derived from household assets and housing characteristics. Wealth quintiles were constructed separately within each survey and therefore represented relative socioeconomic position within each survey rather than absolute changes in living standards over time.

For inequality analyses, children were ranked according to their household wealth score within each survey. The rank was expressed as a weighted fractional position ranging from 0 (poorest) to 1 (richest), permitting comparison of socioeconomic gradients across survey rounds.

Residence was classified as urban or rural according to the definition used in each survey [13–15]. To examine the contribution of the capital-area population to urban trends, urban clusters in Dhaka district were classified separately from other urban areas. For analyses specific to 2025, we additionally distinguished the three sampled city corporations-Dhaka North, Dhaka South, and Chattogram from other urban and rural areas.

Covariates were selected a priori by the investigators based on their potential relevance to child anthropometry and the associations under study. These included child age group (0-5, 6-11, 12-23, 24-35, 36-47, and 48-59 months), sex, maternal education, administrative division, household size, and religion of the household head. Maternal education was harmonised into three categories for pooled analyses and four categories for survey-specific analyses. Administrative divisions were harmonised to the eight-division structure used in 2025; the four districts subsequently incorporated into Mymensingh division were assigned to that division in the 2012-2013 data.

### Statistical analysis

All estimates incorporated the child sampling weights and accounted for survey stratification and clustering. Stratum and cluster identifiers were made unique across survey rounds for pooled analyses. Subgroup estimates were obtained using survey subpopulation procedures to preserve the full survey design. Confidence intervals for proportions were calculated using a logit transformation. Estimates are reported with 95% confidence intervals (CIs), and two-sided P values are reported where available.

### Temporal trends in overweight

We estimated survey-weighted overweight prevalence nationally and by residence, wealth quintile, administrative division, child age group, sex, and maternal education for each survey round. Differences between survey rounds were estimated using survey-weighted linear probability models fitted to pooled data, with survey round as the explanatory variable. Results are expressed as absolute differences in percentage points with 95% CIs. The same approach was used to estimate changes in mean WHZ.

### Wealth and residential inequalities

We assessed socioeconomic inequalities using the slope index of inequality (SII) and relative index of inequality (RII) [19]. The SII quantified the absolute difference in predicted overweight prevalence across the extremes of the fractional wealth rank, whereas the RII quantified the corresponding relative difference using a Poisson model with a log link. Higher values indicated a greater concentration of overweight among wealthier children.

We estimated crude and adjusted inequality measures. Models were adjusted for child age group and sex, and additional models accounted for wealth and residence together, as appropriate, followed by fully adjusted models incorporating the specified covariates. The RII was interpreted under the assumption of a log-linear relationship between wealth rank and overweight prevalence.

To assess departures from linearity, we fitted restricted cubic spline models for wealth rank using knots at the 10th, 50th, and 90th percentiles and examined quintile-specific prevalence. We assessed whether wealth and residential gradients differed across survey rounds by including survey-by-wealth-rank and survey-by-residence interaction terms in pooled models. We also estimated wealth gradients separately in urban and rural populations and examined urban-by-wealth interaction terms within each survey.

Because the 2025 survey used a new census sampling frame and urban classification, changes in the composition of the urban population could have contributed to temporal differences in urban overweight prevalence. We therefore conducted a secondary direct-standardisation analysis, applying survey-specific urban wealth-quintile prevalence estimates to the urban wealth distribution of other survey rounds.

### Correlates of overweight in 2019 and 2025

We fitted modified Poisson regression models with robust variance estimation to estimate crude and adjusted prevalence ratios (PRs) with 95% CIs for overweight in 2019 and 2025 [20]. The models included child age group, sex, maternal education, household wealth quintile, residence, administrative division, household size, and religion of the household head.

For 2025, an additional model replaced the binary urban-rural variable with a three-category classification of city corporations, other urban areas, and rural areas. We also estimated the proportion of overweight children who were concurrently stunted, overall and by wealth quintile and residence.

These analyses were intended to describe adjusted associations between measured characteristics and overweight. The adjusted PRs were not interpreted as causal effects, because the cross-sectional design and covariate adjustment strategy did not establish causal relationships or necessarily eliminate residual confounding.

### Anthropometric measurement quality and sensitivity analyses

We examined anthropometric measurement quality because overweight was defined by the upper tail of the WHZ distribution. Indicators included the proportion of measured children with implausible WHZ values, the mean, standard deviation (SD), and skewness of WHZ, the proportion measured in the correct position for age, and terminal-digit preference in recorded weight and height measurements.

We also calculated an SD-adjusted overweight prevalence, defined as the expected proportion with WHZ greater than +2 if the observed mean WHZ were retained but the SD were set to 1. This measure was used as a diagnostic indicator of the potential contribution of excess dispersion to observed overweight prevalence, rather than as an estimate of corrected or true prevalence.

Following identification of an unusually high overweight prevalence in urban Dhaka district in 2019, we examined overweight case counts and WHZ dispersion by measurer in that area. Analyses prompted by this finding were considered post hoc. We repeated the principal trend and inequality analyses after excluding urban Dhaka district to assess the influence of this area on the findings.

Additional sensitivity analyses examined alternative outcome definitions: body mass index-for-age z-score greater than +2; WHZ greater than +1; WHZ greater than +2 restricted to children aged 6-59 months; WHZ greater than +2 among children measured in the correct position for age; and overweight without concurrent stunting. These analyses assessed the sensitivity of the findings to the anthropometric index, threshold, age range, measurement position, and potential misclassification associated with inaccurate length or height measurement.

To assess potential selection bias due to missing or implausible anthropometric measurements, we compared children with and without a valid WHZ and repeated the principal analyses using response-adjusted survey weights. The response-adjusted weight was calculated by multiplying the original survey weight by the inverse of the modelled probability of having a valid WHZ. This probability was estimated separately within each survey using child age group, sex, residence, wealth quintile, administrative division, and maternal education. Four children in the 2025 analytic sample lacked maternal education data and were excluded from fully adjusted models requiring this covariate.

### Statistical software and reporting

All statistical analyses were done in Stata version 17 (StataCorp, College Station, TX, USA). Figures were prepared in R version 4.6.1 using the *ggplot2* and *patchwork* packages, based on statistical analysis outputs. Before conducting the substantive analyses, we compared the estimated national, urban, rural, and wealth-quintile overweight prevalence with the corresponding estimates published in the three MICS survey reports(Supplementary Table S1).

The primary analyses were the survey-weighted prevalence estimates and temporal and socioeconomic inequality measures. Interaction tests and secondary or post hoc analyses were not adjusted for multiple comparisons and were interpreted as exploratory. The study was reported in accordance with the Strengthening the Reporting of Observational Studies in Epidemiology (STROBE) guidance for cross-sectional studies [21].

### Ethics statement

The 2019 survey protocol was approved by a Government of Bangladesh technical committee led by BBS, and the 2025 survey protocol was approved by the icddr,b Ethical Review Committee (PR-24113). Respondents gave verbal consent in each survey. We used anonymised public data obtained from UNICEF with permission. No further ethical approval was needed for this secondary analysis.

## Results

Across the three Bangladesh Multiple Indicator Cluster Surveys (MICS), 72,768 children aged under five years were listed in sampled households, of whom 67,359 had a completed child questionnaire. Anthropometric measurements were unavailable for 2,840 children, and 547 children with recorded measurements had biologically implausible weight-for-height z-scores (WHZ) and were excluded. The final analytical sample comprised 63,972 children: 19,598 in 2012-13, 22,063 in 2019, and 22,311 in 2025. These represented 95.0% of children with a completed questionnaire.

Among children with recorded anthropometry, 0.8% to 0.9% were excluded because of implausible WHZ values. Children without valid WHZ measurements were more likely to be younger than six months and, in 2019, to live in urban areas and belong to the richest wealth quintile (Supplementary Table S9).

The age and sex distributions were broadly similar across surveys (Table 1). Boys accounted for 51.2%, 52.0%, and 51.8% of children in 2012-13, 2019, and 2025, respectively. Maternal education increased over time: the proportion whose mothers had no formal schooling or pre-primary education declined from 22.3% in 2012-13 to 6.0% in 2025, while the proportion whose mothers had secondary or higher education increased from 47.9% to 76.2%. The weighted proportion of children living in urban areas increased from 20.4% to 28.7%.

**Table 1.** Characteristics of children aged 0-59 months with valid weight-for-height, Bangladesh MICS 2012-13, 2019 and 2025.

| Characteristic | Category | 2012-13<br>(n=19,598) | 2019 (n=22,063) | 2025 (n=22,311) |
| --- | --- | --- | --- | --- |
| Residence | Rural | 16,472 (79.6) | 18,020 (79.2) | 16,392 (71.3) |
|  | Urban | 3,126 (20.4) | 4,043 (20.8) | 5,919 (28.7) |
| Wealth quintile | Poorest | 5,871 (24.5) | 5,479 (21.8) | 5,221 (20.0) |
|  | Second | 4,231 (20.4) | 4,628 (19.7) | 4,776 (19.9) |
|  | Middle | 3,562 (18.7) | 4,188 (18.8) | 4,455 (19.9) |
|  | Fourth | 3,198 (18.0) | 4,133 (19.6) | 4,216 (20.9) |
|  | Richest | 2,736 (18.4) | 3,635 (20.1) | 3,643 (19.3) |
| **Mother's education** | None or pre-primary | 4,886 (22.3) | 2,457 (11.1) | 1,395 (6.0) |
|  | Primary | 5,882 (29.8) | 5,320 (23.7) | 4,026 (17.9) |
|  | Secondary or higher | 8,830 (47.9) | 14,286 (65.2) | 16,886 (76.2) |
| **Sex** | Male | 10,065 (51.2) | 11,411 (52.0) | 11,514 (51.8) |
|  | Female | 9,533 (48.8) | 10,652 (48.0) | 10,797 (48.2) |
| **Age (months)** | 0-5 | 1,809 (9.3) | 2,195 (10.1) | 2,071 (9.4) |
|  | 6-11 | 1,874 (9.8) | 2,133 (9.8) | 2,168 (9.8) |
|  | 12-23 | 3,860 (20.0) | 4,389 (19.6) | 4,649 (21.0) |
|  | 24-35 | 3,842 (19.6) | 4,329 (19.6) | 4,640 (21.0) |
|  | 36-47 | 4,073 (20.6) | 4,568 (20.8) | 4,470 (19.7) |
|  | 48-59 | 4,140 (20.7) | 4,449 (20.1) | 4,313 (19.1) |
| **Division** | Barishal | 1,846 (6.2) | 1,999 (5.8) | 1,894 (5.4) |
|  | Chattogram | 4,048 (23.2) | 4,532 (21.4) | 4,904 (23.4) |
|  | Dhaka | 3,676 (22.7) | 4,349 (23.8) | 4,689 (24.3) |
|  | Khulna | 2,612 (9.8) | 3,071 (10.6) | 2,987 (10.3) |
|  | Mymensingh | 1,256 (8.0) | 1,328 (7.6) | 1,331 (8.0) |
|  | Rajshahi | 1,899 (11.6) | 2,325 (12.1) | 2,260 (11.6) |
|  | Rangpur | 2,483 (11.3) | 2,615 (10.8) | 2,705 (10.8) |
|  | Sylhet | 1,778 (7.1) | 1,844 (7.9) | 1,541 (6.3) |
| **Household members** | 1-4 | 7,059 (37.2) | 8,777 (40.2) | 8,564 (39.9) |
|  | 5-6 | 7,657 (37.6) | 8,625 (38.6) | 9,208 (39.9) |
|  | 7 or more | 4,882 (25.2) | 4,661 (21.2) | 4,539 (20.1) |
| **Religion of household head** | Other religion | 2,218 (8.7) | 2,198 (8.4) | 1,923 (7.0) |
|  | Islam | 17,380 (91.3) | 19,865 (91.6) | 20,388 (93.0) |

*Values are unweighted numbers (weighted percentages). Percentages may not sum to 100* *because of rounding.*

### Prevalence trends and anthropometric data quality

National overweight prevalence increased from 1.6% (95% CI 1.4-1.9) in 2012-13 to 2.4% (2.2-2.7) in 2019, before declining to 1.7% (1.5-2.0) in 2025 (Table 2; Figure 1). The pattern differed by residence. Urban prevalence was 2.7%, 4.8%, and 1.7%, respectively, while rural prevalence was 1.3%, 1.8%, and 1.7%. Prevalence was consistently higher in the richest than in the poorest wealth quintile: 3.4% versus 0.9% in 2012-13, 5.4% versus 1.5% in 2019, and 2.6% versus 0.9% in 2025. Prevalence was highest in infancy in all three surveys, at 3.0% to 4.2% among children aged 0-5 months. Dhaka division had the highest prevalence in 2019 (4.7%) and 2025 (3.4%), and Sylhet division in 2012-13 (2.9%).

**Fig 1.**
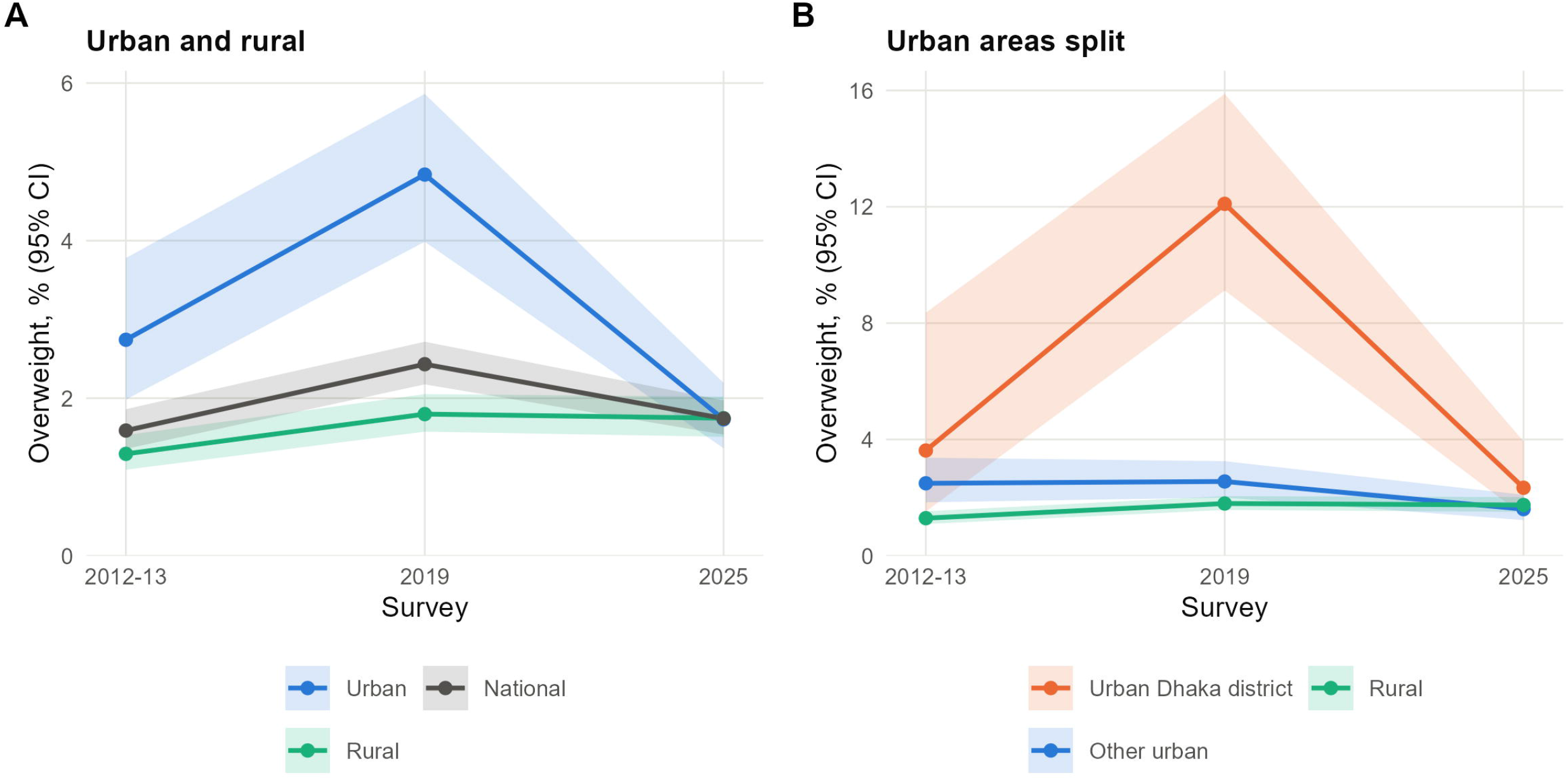
Trend in overweight by place of residence, children aged 0 to 59 months, Bangladesh MICS 2012-13, 2019 and 2025. (A) Urban, rural and national prevalence. (B) Urban areas split into urban Dhaka district and other urban areas, with rural areas for comparison. Urban Dhaka district covers urban clusters of Dhaka district, including its city corporations. Lines show survey-weighted prevalence and shaded bands show 95% confidence intervals. Overweight is WHZ above +2 (WHO 2006).

**Table 2.** Prevalence of overweight (WHZ >+2) in children aged 0-59 months, Bangladesh MICS 2012-13, 2019 and 2025.

| Group | Category | 2012-13 %<br>(95% CI) | 2019 % (95%<br>CI) | 2025 % (95%<br>CI) |
| --- | --- | --- | --- | --- |
| **Total** |  | 1.6 (1.4-1.9) | 2.4 (2.2-2.7) | 1.7 (1.5-2.0) |
| **Residence** | Urban | 2.7 (2.0-3.8) | 4.8 (4.0-5.9) | 1.7 (1.4-2.2) |
|  | Rural | 1.3 (1.1-1.5) | 1.8 (1.6-2.1) | 1.7 (1.5-2.0) |
|  | Urban Dhaka district | 3.6 (1.5-8.4) | 12.1 (9.1-15.9) | 2.3 (1.4-3.9) |
|  | Other urban | 2.5 (1.8-3.4) | 2.6 (2.0-3.3) | 1.6 (1.2-2.1) |
| **Wealth quintile** | Poorest | 0.9 (0.7-1.2) | 1.5 (1.2-2.0) | 0.9 (0.6-1.3) |
|  | Second | 1.2 (0.9-1.6) | 1.3 (0.9-1.7) | 1.3 (1.0-1.8) |
|  | Middle | 1.2 (0.9-1.8) | 1.7 (1.3-2.2) | 1.8 (1.3-2.3) |
|  | Fourth | 1.5 (1.1-2.1) | 2.3 (1.8-2.9) | 2.1 (1.7-2.7) |
|  | Richest | 3.4 (2.5-4.6) | 5.4 (4.5-6.4) | 2.6 (2.1-3.3) |
| **Mother's education** | None or pre-primary | 1.1 (0.8-1.5) | 1.6 (1.1-2.2) | 1.9 (1.1-3.2) |
|  | Primary | 1.5 (1.1-2.0) | 1.9 (1.5-2.5) | 1.4 (1.0-1.9) |
|  | Secondary or higher | 1.9 (1.5-2.4) | 2.8 (2.4-3.1) | 1.8 (1.6-2.1) |
| **Age (months)** | 0-5 | 4.1 (2.9-5.8) | 4.2 (3.3-5.3) | 3.0 (2.3-3.9) |
|  | 6-11 | 1.5 (1.0-2.2) | 3.2 (2.4-4.2) | 3.0 (2.3-4.1) |
|  | 12-23 | 1.2 (0.9-1.8) | 2.7 (2.2-3.4) | 1.7 (1.3-2.3) |
|  | 24-35 | 1.2 (0.8-1.6) | 2.4 (1.8-3.1) | 1.2 (0.9-1.7) |
|  | 36-47 | 1.6 (1.1-2.4) | 2.0 (1.5-2.5) | 1.5 (1.1-2.0) |
|  | 48-59 | 1.3 (0.9-1.8) | 1.4 (1.0-1.9) | 1.2 (0.9-1.7) |
| **Sex** | Male | 1.6 (1.3-2.0) | 2.6 (2.3-3.0) | 1.8 (1.6-2.2) |
|  | Female | 1.6 (1.2-2.0) | 2.3 (1.9-2.7) | 1.6 (1.4-1.9) |
| **Division** | Barishal | 0.8 (0.5-1.4) | 1.9 (1.3-2.9) | 1.4 (0.9-2.0) |
|  | Chattogram | 1.2 (0.9-1.7) | 1.8 (1.4-2.3) | 1.2 (0.9-1.7) |
|  | Dhaka | 2.5 (1.8-3.4) | 4.7 (3.9-5.6) | 3.4 (2.9-4.1) |
|  | Khulna | 1.1 (0.7-1.7) | 1.3 (0.9-2.0) | 1.1 (0.7-1.6) |
|  | Mymensingh | 1.4 (0.9-2.2) | 1.6 (1.0-2.6) | 1.3 (0.7-2.5) |
|  | Rajshahi | 1.2 (0.7-2.1) | 1.8 (1.3-2.5) | 1.2 (0.8-1.9) |
|  | Rangpur | 1.1 (0.6-2.0) | 2.4 (1.8-3.2) | 0.9 (0.6-1.3) |
|  | Sylhet | 2.9 (2.0-4.2) | 1.0 (0.6-1.7) | 1.6 (1.0-2.7) |

*Estimates are survey weighted. Overweight was defined as WHZ > +2 according to the 2006* *WHO Child Growth Standards.*

The 2019 peak was concentrated in urban Dhaka district, where prevalence reached 12.1% (95% CI 9.1-15.9), compared with 3.6% in 2012-13 and 2.3% in 2025 (Supplementary Table S6). This area accounted for 5.0% of measured children. Measurement patterns warrant caution: one measurer assessed 508 of the 526 children measured in urban Dhaka district in 2019 and recorded all 67 overweight cases. WHZ variability was higher in urban Dhaka district (SD 1.66) than in other urban areas (1.20) or rural areas (1.17). After excluding urban Dhaka district, national prevalence was 1.5%, 1.9%, and 1.7% across the three surveys. The difference between 2019 and 2025 was not statistically significant (-0.21 percentage points, 95% CI -0.53 to 0.10; p=0.180).

The decline in overweight prevalence between 2019 and 2025 coincided with a decrease in mean WHZ of 0.16 z-score units (95% CI -0.19 to -0.13) and an increase in wasting prevalence from 9.8% to 12.9% (Supplementary Tables S2-S3). Mean WHZ declined across all wealth quintiles. Observed overweight prevalence exceeded the corresponding SD-adjusted estimates in every survey; in 2012-13, observed prevalence was 1.6%, compared with an SD-adjusted estimate of 0.35%. WHZ variability was highest in 2019 (SD 1.22), particularly among urban and richest-quintile children. Analyses using alternative anthropometric definitions showed a broadly similar temporal pattern (Supplementary Tables S4 and S8).

### Wealth and residential inequalities

Overweight was consistently concentrated among wealthier children (Table 3A; Figures 2 and 3). The crude relative index of inequality (RII) was 5.87 (95% CI 3.11-11.09) in 2012-13, 7.05 (4.37-11.39) in 2019, and 3.72 (2.40-5.77) in 2025. The fully adjusted RII estimates were 3.66 (1.93-6.93), 3.55 (2.21-5.71), and 3.35 (2.02-5.54), respectively, with no evidence of a temporal change in the adjusted wealth gradient (interaction p=0.974). The fully adjusted slope index of inequality (SII) was 1.9 percentage points (95% CI 1.0-2.9) in 2012-13, 2.7 (1.8-3.7) in 2019, and 2.0 (1.1-2.8) in 2025.

**Fig 2.**
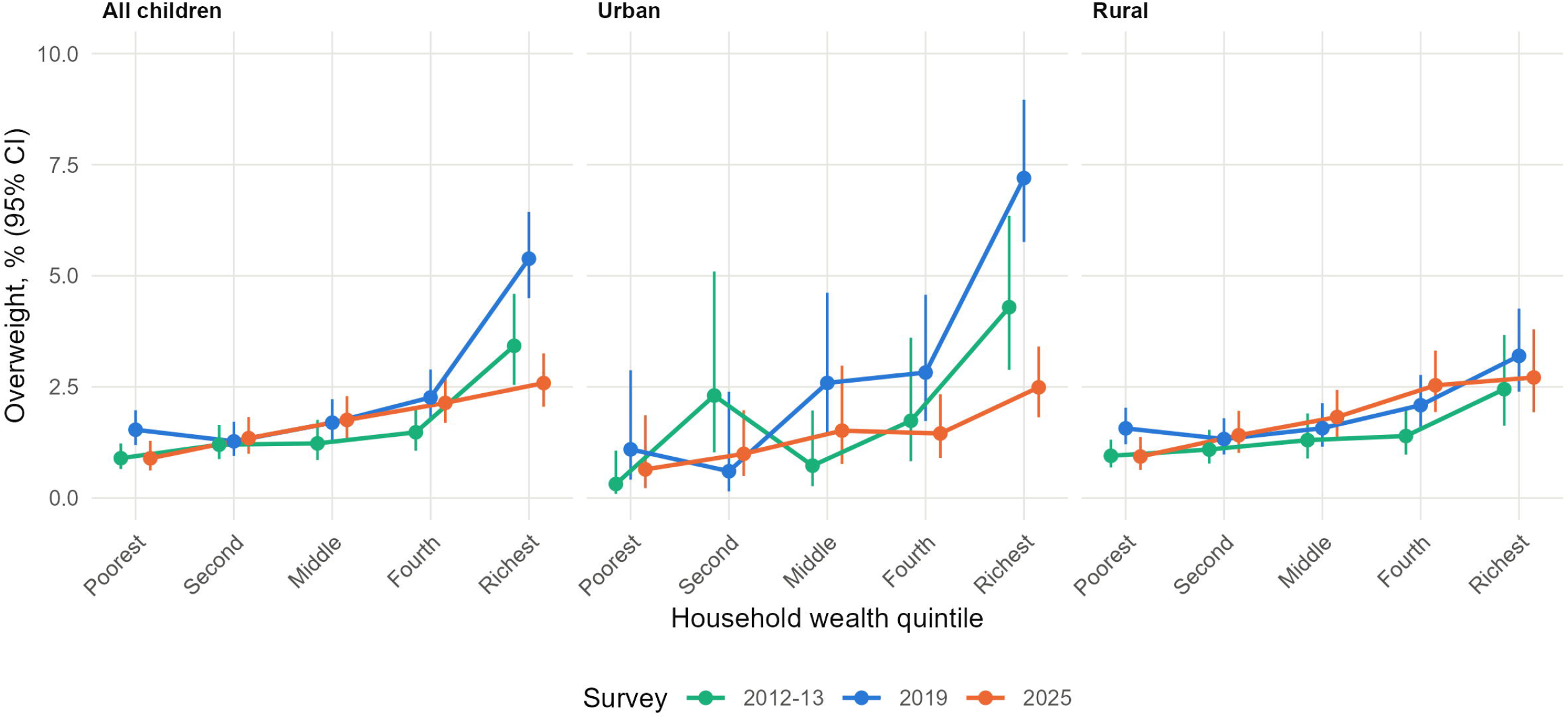
Overweight across household wealth quintiles in each survey. Panels show all children, urban children and rural children. Points show survey-weighted prevalence and bars show 95% confidence intervals. The vertical axis is truncated at 10%.

**Fig 3.**
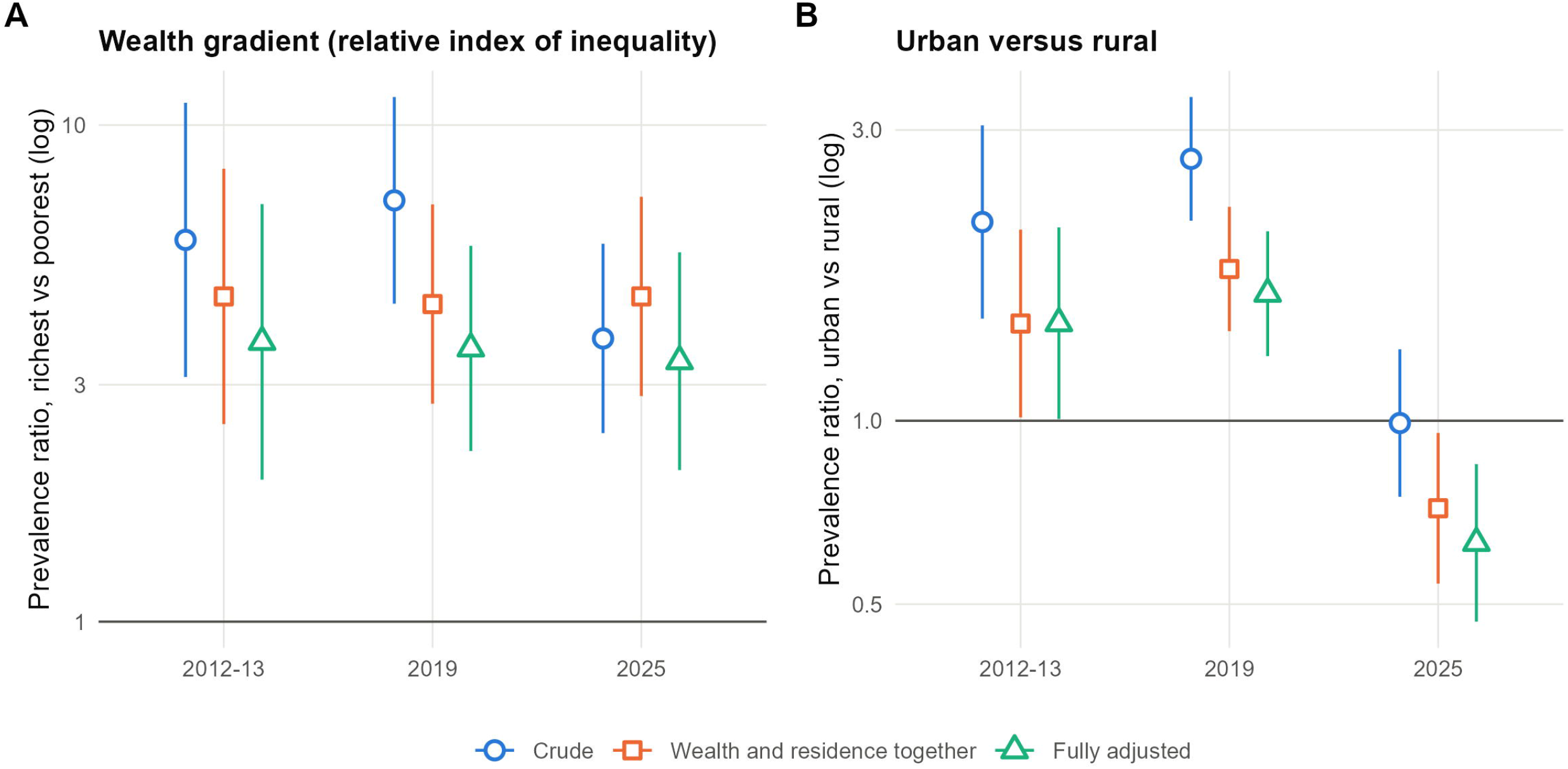
Wealth and urban gradients in overweight, crude and adjusted. (A) Relative index of inequality, the prevalence ratio between the top and bottom of the wealth rank. (B) Prevalence ratio for urban against rural residence. Full adjustment covers age group, sex, mother’s education and division, with residence or wealth rank as relevant. Bars show 95% confidence intervals on a log scale.

**Table 3A.** Wealth gradient: SII and RII (Bangladesh MICS)

| Measure (95% CI) | 2012-13 | 2019 | 2025 |
| --- | --- | --- | --- |
| SII, crude (% points) | 2.7 (1.6-3.8) | 4.5 (3.3-5.6) | 2.2 (1.5-3.0) |
| SII, fully adjusted (% points) | 1.9 (1.0-2.9) | 2.7 (1.8-3.7) | 2.0 (1.1-2.8) |
| RII, crude | 5.87 (3.11-11.09) | 7.05 (4.37-11.39) | 3.72 (2.40-5.77) |
| RII, adj. age/sex | 5.84 (3.11-10.98) | 6.98 (4.31-11.29) | 3.59 (2.31-5.58) |
| RII, adj. residence | 4.52 (2.50-8.17) | 4.36 (2.75-6.92) | 4.52 (2.85-7.17) |
| RII, fully adjusted | 3.66 (1.93-6.93) | 3.55 (2.21-5.71) | 3.35 (2.02-5.54) |
| RII (urban only), crude | 19.34 (3.35-111.63) | 27.34 (7.88-94.82) | 6.13 (2.10-17.92) |
| SII (urban only), crude (% points) | 5.2 (2.4-7.9) | 9.8 (6.9-12.8) | 2.5 (1.2-3.9) |
| RII (rural only), crude | 3.00 (1.54-5.85) | 2.45 (1.41-4.24) | 4.14 (2.47-6.96) |
| SII (rural only), crude (% points) | 1.4 (0.5-2.4) | 1.6 (0.6-2.6) | 2.5 (1.5-3.5) |
| Mean WHZ diff (top vs bottom wealth) | 0.45 (0.36-0.54) | 0.57 (0.50-0.65) | 0.45 (0.38-0.52) |
| Non-linearity *p* (crude; fully adj) | 0.002; 0.016 | <0.001; <0.001 | 0.675; 0.508 |

*Estimates are shown with 95% CIs unless otherwise indicated. SII, slope index of inequality; RII, relative index of inequality; WHZ, weight-for-height z-score. Minimal adjustment covers age group and sex. Full adjustment covers age group, sex, maternal education and division, with residence or wealth rank as relevant. Non-linearity *p* values test the second spline term of wealth rank. Wealth-rank interaction: *p*=0.140 (crude), *p*=0.974 (fully adjusted). Urban-by-survey interaction: *p*<0.001 (crude and fully adjusted). Urban-by-wealth-rank interaction: *p*=0.052, *p*<0.001, and *p*=0.519 for 2012-13, 2019, and 2025, respectively. Overweight cases / children: 303/19,598; 456/22,063; and 368/22,311. Fully adjusted 2025 models exclude four children with missing maternal education.*

The shape of the wealth gradient differed across surveys. In 2012-13 and 2019, overweight was disproportionately concentrated at the wealthiest end of the distribution, with evidence of non-linearity in crude models (p=0.002 and p<0.001, respectively). In 2025, the gradient was more consistent with a linear relationship (p=0.675).

The association between urban residence and overweight changed over time (urban-by-survey interaction p<0.001; Table 3B). The crude urban-to-rural prevalence ratio was 2.12 (95% CI 1.47-3.05) in 2012-13, 2.69 (2.13-3.40) in 2019, and 0.99 (0.75-1.31) in 2025. Adjustment for age and sex did not materially change these estimates. After adjustment for wealth rank, prevalence ratios were 1.44 (1.01-2.06), 1.77 (1.40-2.24), and 0.72 (0.54-0.95), respectively. In the fully adjusted model, the corresponding estimates were 1.45 (1.01-2.08), 1.62 (1.28-2.05), and 0.63 (0.47-0.85). The lower adjusted urban prevalence in 2025 persisted after excluding urban Dhaka district.

**Table 3B.** Urban vs Rural - Prevalence Ratios (Bangladesh MICS)

| Measure (95% CI) | 2012-13 | 2019 | 2025 |
| --- | --- | --- | --- |
| PR (urban vs rural, crude) | 2.12 (1.47-3.05) | 2.69 (2.13-3.40) | 0.99 (0.75-1.31) |
| PR (adj. age/sex) | 2.15 (1.50-3.10) | 2.68 (2.12-3.39) | 1.00 (0.76-1.32) |
| PR (adj. wealth) | 1.44 (1.01-2.06) | 1.77 (1.40-2.24) | 0.72 (0.54-0.95) |
| PR (fully adjusted) | 1.45 (1.01-2.08) | 1.62 (1.28-2.05) | 0.63 (0.47-0.85) |

In 2025, the absolute wealth gradient was similar in urban and rural areas, with an SII of 2.5 percentage points in each group; the urban-by-wealth interaction was not statistically significant (p=0.519). The proportion of urban children in the richest wealth quintile declined from 52.8% in 2019 to 38.3% in 2025. Applying the 2025 wealth-quintile-specific prevalence estimates to the 2019 urban wealth distribution yielded an estimated prevalence of 1.9%, compared with the observed 2025 prevalence of 1.7%. Within the richest urban quintile, prevalence declined from 7.2% in 2019 to 2.5% in 2025.

### Correlates of overweight and concurrent stunting

In survey-specific adjusted models, overweight prevalence in 2025 was highest among infants and declined with age (Figure 4; Supplementary Table S5). Prevalence was 3.0% among children aged 0-11 months and 1.2% among those aged 48-59 months. Compared with infants aged 0-5 months, children aged 48-59 months had a lower adjusted prevalence (PR 0.46, 95% CI 0.30-0.69).

**Fig 4.**
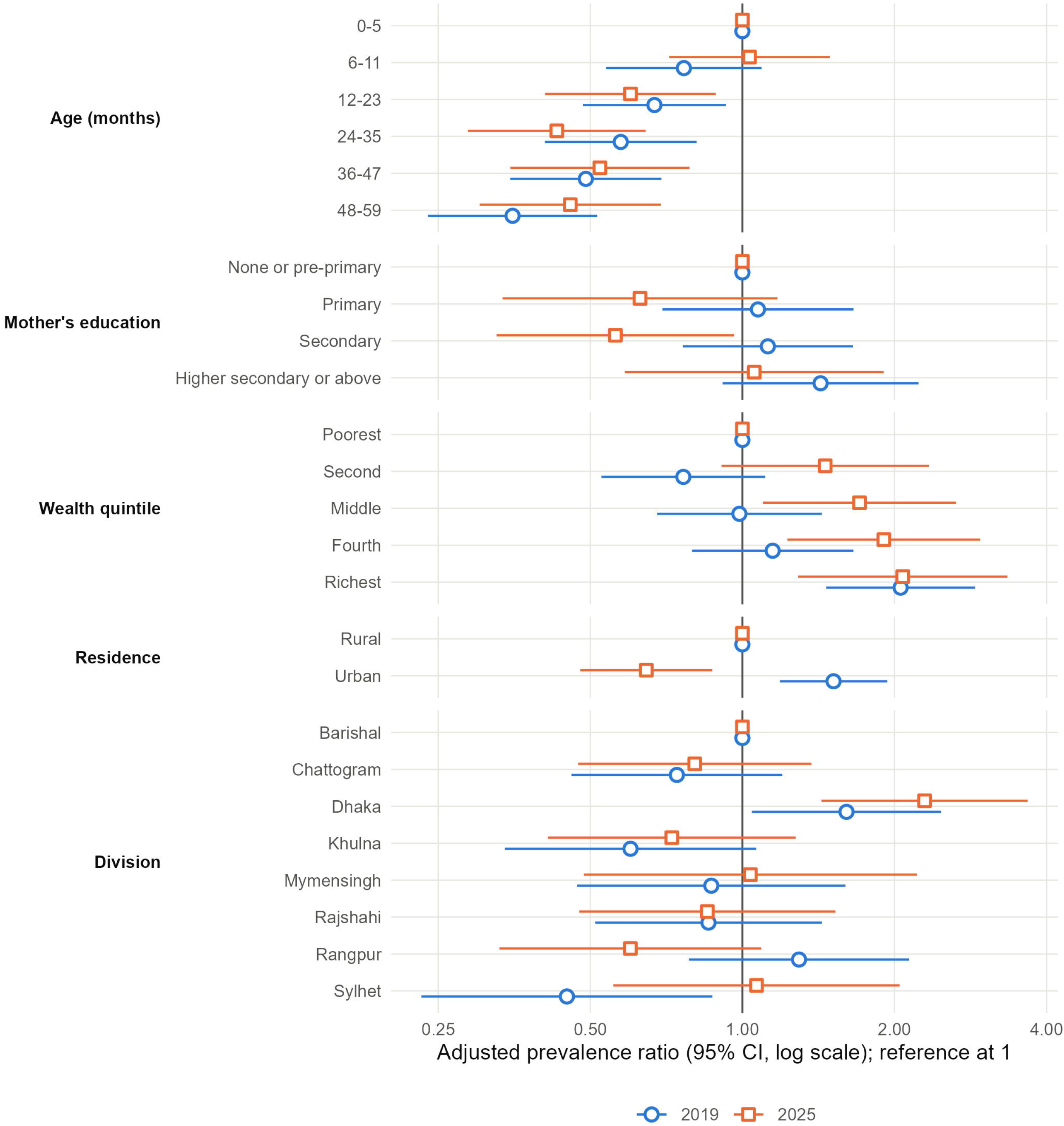
Adjusted prevalence ratios for overweight, 2019 and 2025. Modified Poisson models including age group, sex, mother’s education, wealth quintile, residence, division, household size and religion. Points at 1 are reference categories. Each ratio is adjusted for the other variables shown and describes an independent correlate; it is not adjusted for its own confounders. Bars show 95% confidence intervals on a log scale.

Household wealth was associated with overweight in 2025. Compared with children in the poorest quintile, adjusted PRs were 1.71 (95% CI 1.10-2.65) in the middle quintile, 1.91 (1.23-2.96) in the fourth quintile, and 2.08 (1.29-3.35) in the richest quintile. In 2019, the association was more concentrated in the richest quintile, which had an adjusted PR of 2.06 (1.47-2.89) relative to the poorest quintile.

In these survey-specific models, urban residence was associated with higher overweight prevalence in 2019 (adjusted PR 1.52, 95% CI 1.19-1.93) and lower prevalence in 2025 (0.65, 0.48-0.87). In the 2025 three-category residence model, adjusted PRs were 0.46 (0.27-0.78) for city corporations and 0.72 (0.53-0.98) for other urban areas, compared with rural areas. Children in Dhaka division had a higher adjusted prevalence than those in Barishal division (PR 2.29, 1.43-3.67). Maternal education showed no consistent adjusted gradient, and there was little evidence of independent associations with sex, household size, or religion of the household head.

In 2025, 41.4% (95% CI 35.3-47.7) of overweight children were also stunted (Supplementary Table S7). This proportion was higher in the poorest than in the richest wealth quintile (57.9% versus 26.7%). Overweight without concurrent stunting showed a steeper wealth gradient than overweight overall (RII 9.61, 95% CI 5.00-18.48). The wealth-adjusted urban-to-rural prevalence ratio for overweight without concurrent stunting was 0.71 (0.49-1.03).

### Sensitivity analyses

The principal findings were similar in analyses using response-adjusted survey weights to account for missing or implausible anthropometric measurements (Supplementary Table S10). National overweight prevalence remained 1.6% in 2012-13, 2.4% in 2019, and 1.7% in 2025. The fully adjusted RII estimates were 3.59, 3.52, and 3.34, respectively, and the wealth-adjusted urban-to-rural prevalence ratios were 1.44, 1.76, and 0.72. Analyses excluding urban Dhaka district and using alternative anthropometric definitions are reported in Supplementary Tables S4 and S8.

## Discussion

In this analysis of three nationally representative surveys, overweight among children younger than 5 years in Bangladesh remained uncommon, increasing from 1.6% in 2012-2013 to 2.4% in 2019 before declining to 1.7% in 2025. However, this overall pattern concealed persistent wealth inequalities and a changing association with urban residence. Interpretation of the temporal trend requires caution because the 2019 increase was concentrated in urban Dhaka district, where anthropometric measurement patterns raised concerns about data quality.

The national estimates are broadly consistent with previous analyses reporting low overweight prevalence among Bangladeshi children under five [7,8]. However, differences in survey populations, definitions and analytical methods limit direct comparisons. The decline after 2019 also coincided with a reduction in mean weight-for-height z-score (WHZ) and an increase in wasting prevalence from 9.8% to 12.9%. Thus, lower overweight prevalence should not necessarily be interpreted as improved nutritional status. These concurrent changes highlight the importance of assessing overweight alongside indicators of undernutrition [2,3].

The 2019 peak warrants particular caution. Overweight prevalence reached 12.1% in urban Dhaka district, where one measurer assessed 508 of 526 children and recorded all 67 overweight cases. WHZ variability was also greater than in other urban and rural areas. Because random measurement error can increase the dispersion of anthropometric measurements and affect classification around thresholds, these findings raise concerns about the reliability of the local estimate [10–12]. Measurer and location were strongly confounded, so the data cannot distinguish a measurement-related excess from a genuine local difference. After excluding urban Dhaka district, national prevalence was 1.5%, 1.9% and 1.7% across the three surveys, and the difference between 2019 and 2025 was not statistically significant. The apparent national peak should therefore be interpreted cautiously rather than assumed to represent a true population-wide increase.

Household wealth was consistently associated with overweight. The fully adjusted relative index of inequality (RII) was 3.66 (95% CI 1.93-6.93) in 2012-2013, 3.55 (2.21-5.71) in 2019 and 3.35 (2.02-5.54) in 2025, with no evidence of temporal change (interaction p=0.974). The fully adjusted slope index of inequality (SII) was 1.9, 2.7 and 2.0 percentage points, respectively. This finding is consistent with evidence of socioeconomic inequalities in childhood malnutrition across South Asia [6] and the broader double burden of malnutrition in low- and middle-income countries [2,3]. Although the relative wealth gradient persisted, overweight remained uncommon and absolute differences were comparatively modest. Both absolute and relative measures are therefore important for interpreting the public health significance of these inequalities. The shape of the gradient also differed across surveys: the concentration at the wealthiest end was more pronounced in earlier rounds, whereas the 2025 pattern was more graded. This difference does not establish that overweight has spread to middle-income households over time, because wealth quintiles represent relative socioeconomic positions within each survey. In 2025, the absolute wealth gradient was also similar in urban and rural areas (2.5 percentage points in each), so wealth-related overweight was not confined to cities.

The association between urban residence and overweight changed substantially across surveys. The wealth-adjusted urban-to-rural prevalence ratio was 1.44 (95% CI 1.01-2.06) in 2012-2013 and 1.77 (1.40-2.24) in 2019 but declined to 0.72 (0.54-0.95) in 2025. This reversal should not be interpreted as evidence that urban residence is protective. The 2025 survey used a new census frame and urban-rural classification, potentially affecting comparability with earlier rounds [15]. Although the decline in the proportion of urban children belonging to the richest quintile explained little of the observed difference in the standardisation analysis, changes in population composition, measurement conditions and unmeasured contextual factors remain possible explanations. Further research using comparable geographic classifications is needed to establish whether this reversal persists.

In 2025, 41.4% of children classified as overweight were also stunted, with a higher proportion among children in the poorest than the richest quintile (57.9% versus 26.7%). This signifies the double burden of malnutrition and reinforces the importance of considering multiple dimensions of nutritional status rather than treating overweight and undernutrition as mutually exclusive [2,3,6,22]. Overweight prevalence was also highest in infancy, although the repeated cross-sectional design cannot establish individual growth trajectories or explain this age pattern.

### Implications

National programmes do not need to treat overweight in children under five as an emerging epidemic. They do need to watch wealthier families in both cities and villages, where early overweight persists. Infant and young child feeding counselling could add advice on sugary drinks and processed snacks for these families. Survey programmes should report the SD of WHZ and SD-adjusted prevalence with every overweight estimate. Measurer-level standardisation and field checks would make trends in this rare outcome easier to interpret.

### Strengths and limitations

This study provides a nationally representative assessment of childhood overweight in Bangladesh across three MICS rounds spanning 2012-13 to 2025. Its main strengths include the use of a consistent analytical framework, examination of both absolute and relative socioeconomic inequalities, and assessment of wealth and residence as distinct dimensions of inequality. Sensitivity analyses addressing missing anthropometric measurements and the influence of urban Dhaka district further strengthened the assessment of the robustness of the findings.

The findings should nevertheless be interpreted in light of several limitations. Approximately 5% of interviewed children lacked valid anthropometric measurements; although response weighting did not materially change the estimates, selection bias cannot be excluded. Comparisons across rounds may also be affected by the new sampling frame and urban-rural classification introduced in 2025. Because wealth quintiles are defined relative to each survey population, they do not measure changes in absolute household living standards. In urban Dhaka district in 2019, measurer and location were confounded, preventing a definitive distinction between measurement-related and genuine local differences. The absence of maternal body mass index and limited dietary information restricted assessment of potential explanatory factors. Finally, the small number of overweight cases reduced precision for some subgroup estimates, and the cross-sectional design precluded causal inference and assessment of individual growth trajectories.

### Conclusions

Overweight among children under five in Bangladesh remained uncommon across the 2012-2013, 2019 and 2025 surveys, with no sustained national increase over the study period. The persistent association with household wealth indicates that socioeconomic inequalities remain relevant, whereas the association with urban residence changed over time and was reversed after adjustment for wealth in 2025. The unusually high prevalence in urban Dhaka district in 2019 warrants caution because of concerns about anthropometric measurement, although a measurement effect cannot be established conclusively. The coexistence of overweight and stunting further underscores the need to assess multiple dimensions of child nutritional status. Continued surveillance should therefore examine socioeconomic inequalities alongside anthropometric data quality, while future research should investigate the factors underlying the observed temporal and geographic differences.

## Supporting information

S1 File. Supplementary tables S1 to S10. They cover reproduction of the survey reports, change between surveys, measurement quality and alternative definitions. They also give full prevalence ratio models, urban Dhaka district results, concurrent overweight and stunting, and a sensitivity analysis excluding urban Dhaka district. S9 and S10 Tables give the participant flow, a comparison of children with and without a valid measurement, and response-weighted estimates.

## Declarations

## Data availability

MICS data sets are available from UNICEF on registration at https://mics.unicef.org/surveys. All aggregate results are in the manuscript and S1 File.

