## Supplementary material for "Overweight in children under five in urban and wealthier households in Bangladesh: trends and inequalities across three Multiple Indicator Cluster Surveys, 2012-13 to 2025": Supplement_Draft_v2.docx

S1 File. Supplementary tables

All estimates are survey weighted unless stated otherwise and were produced in Stata 17. Overweight is a weight-for-height z-score (WHZ) above +2 against the WHO 2006 standards.

**S1 Table. Overweight prevalence (%) in the survey reports and in this study.**

Report values are from the nutritional status tables of each survey report (2012-13 Table NU.1; 2019 Table TC.7.1; 2025 Table TC.8.1).

| Group | 2012-13 report | 2012-13 this study | 2019 report | 2019 this study | 2025 report | 2025 this study |
| --- | --- | --- | --- | --- | --- | --- |
| Total | 1.6 | 1.6 | 2.4 | 2.4 | 1.7 | 1.7 |
| Urban | 2.7 | 2.7 | 4.8 | 4.8 | 1.7 | 1.7 |
| Rural | 1.3 | 1.3 | 1.8 | 1.8 | 1.7 | 1.7 |
| Poorest | 0.9 | 0.9 | 1.5 | 1.5 | 0.9 | 0.9 |
| Second | 1.2 | 1.2 | 1.3 | 1.3 | 1.3 | 1.3 |
| Middle | 1.2 | 1.2 | 1.7 | 1.7 | 1.8 | 1.8 |
| Fourth | 1.5 | 1.5 | 2.3 | 2.3 | 2.1 | 2.1 |
| Richest | 3.4 | 3.4 | 5.4 | 5.4 | 2.6 | 2.6 |

**S2 Table. Change in overweight and mean WHZ between surveys.**

Differences from survey-weighted linear models on the pooled data.

Overweight, change in percentage points (95% CI)

| Group | 2019 vs 2012-13 | 2025 vs 2012-13 | 2025 vs 2019 |
| --- | --- | --- | --- |
| Total | 0.84 (0.47 to 1.21); p < 0.001 | 0.15 (-0.18 to 0.48); p = 0.366 | -0.69 (-1.04 to -0.34); p < 0.001 |
| Urban | 2.10 (0.81 to 3.38); p = 0.001 | -1.01 (-1.99 to -0.03); p = 0.042 | -3.11 (-4.13 to -2.09); p < 0.001 |
| Rural | 0.50 (0.18 to 0.83); p = 0.002 | 0.45 (0.12 to 0.79); p = 0.008 | -0.05 (-0.40 to 0.30); p = 0.771 |
| Poorest | 0.64 (0.16 to 1.12); p = 0.009 | -0.00 (-0.43 to 0.43); p = 0.992 | -0.64 (-1.15 to -0.14); p = 0.013 |
| Second | 0.08 (-0.46 to 0.61); p = 0.782 | 0.15 (-0.41 to 0.70); p = 0.603 | 0.07 (-0.48 to 0.63); p = 0.800 |
| Middle | 0.47 (-0.17 to 1.11); p = 0.152 | 0.53 (-0.12 to 1.17); p = 0.108 | 0.06 (-0.60 to 0.72); p = 0.855 |
| Fourth | 0.78 (0.05 to 1.52); p = 0.037 | 0.66 (-0.04 to 1.36); p = 0.064 | -0.12 (-0.87 to 0.63); p = 0.747 |
| Richest | 1.96 (0.56 to 3.36); p = 0.006 | -0.84 (-2.01 to 0.34); p = 0.162 | -2.80 (-3.93 to -1.66); p < 0.001 |
| Urban Dhaka district | 8.48 (3.98 to 12.98); p < 0.001 | -1.28 (-4.53 to 1.97); p = 0.439 | -9.76 (-13.31 to -6.21); p < 0.001 |

Mean weight-for-height z-score, change in z units (95% CI)

| Group | 2019 vs 2012-13 | 2025 vs 2012-13 | 2025 vs 2019 |
| --- | --- | --- | --- |
| Total | 0.06 (0.03 to 0.09); p < 0.001 | -0.09 (-0.12 to -0.06); p < 0.001 | -0.16 (-0.19 to -0.13); p < 0.001 |
| Urban | 0.19 (0.10 to 0.27); p < 0.001 | -0.14 (-0.22 to -0.06); p < 0.001 | -0.33 (-0.39 to -0.26); p < 0.001 |
| Rural | 0.03 (-0.00 to 0.06); p = 0.075 | -0.09 (-0.12 to -0.06); p < 0.001 | -0.12 (-0.15 to -0.09); p < 0.001 |
| Poorest | 0.02 (-0.03 to 0.07); p = 0.434 | -0.11 (-0.16 to -0.06); p < 0.001 | -0.13 (-0.19 to -0.08); p < 0.001 |
| Second | 0.01 (-0.05 to 0.06); p = 0.782 | -0.11 (-0.17 to -0.06); p < 0.001 | -0.12 (-0.18 to -0.07); p < 0.001 |
| Middle | 0.05 (-0.01 to 0.11); p = 0.106 | -0.08 (-0.14 to -0.02); p = 0.012 | -0.13 (-0.19 to -0.07); p < 0.001 |
| Fourth | 0.11 (0.04 to 0.17); p = 0.001 | -0.06 (-0.13 to 0.00); p = 0.061 | -0.17 (-0.24 to -0.11); p < 0.001 |
| Richest | 0.08 (-0.00 to 0.17); p = 0.065 | -0.15 (-0.23 to -0.06); p < 0.001 | -0.23 (-0.30 to -0.15); p < 0.001 |
| Urban Dhaka district | 0.41 (0.14 to 0.68); p = 0.003 | -0.22 (-0.46 to 0.02); p = 0.076 | -0.62 (-0.81 to -0.44); p < 0.001 |

**S3 Table. Measurement quality indicators and SD-adjusted overweight prevalence.**

SD-adjusted prevalence is the share expected above +2 if the observed mean were kept and the SD were 1 (Mei and Grummer-Strawn 2007).

| Indicator | 2012-13 | 2019 | 2025 |
| --- | --- | --- | --- |
| Children with weight and height recorded | 19,762 | 22,266 | 22,491 |
| Implausible WHZ among measured children, % | 0.8 | 0.9 | 0.8 |
| Mean WHZ | -0.69 | -0.63 | -0.79 |
| SD of WHZ | 1.10 | 1.22 | 1.17 |
| Skewness of WHZ | 0.36 | 0.35 | 0.34 |
| SD of HAZ | 1.41 | 1.37 | 1.28 |
| Correct measurement position, % | 99.3 | 99.5 | 95.5 |
| Heights ending in .0 or .5, % (expected 20) | 25.3 | 14.6 | 13.0 |
| Weights ending in .0 or .5, % (expected 20) | 20.1 | 17.9 | 17.5 |

Mean WHZ / SD of WHZ / observed overweight % / SD-adjusted overweight %

| Group | 2012-13 | 2019 | 2025 |
| --- | --- | --- | --- |
| Total | -0.69 / 1.10 / 1.6 / 0.35 | -0.63 / 1.22 / 2.4 / 0.42 | -0.79 / 1.17 / 1.7 / 0.26 |
| Rural | -0.72 / 1.08 / 1.3 / 0.33 | -0.69 / 1.17 / 1.8 / 0.36 | -0.81 / 1.18 / 1.7 / 0.25 |
| Urban | -0.60 / 1.16 / 2.7 / 0.46 | -0.41 / 1.34 / 4.8 / 0.79 | -0.74 / 1.17 / 1.7 / 0.30 |
| Poorest | -0.82 / 1.06 / 0.9 / 0.24 | -0.80 / 1.16 / 1.5 / 0.26 | -0.93 / 1.09 / 0.9 / 0.17 |
| Second | -0.79 / 1.07 / 1.2 / 0.27 | -0.78 / 1.15 / 1.3 / 0.27 | -0.90 / 1.13 / 1.3 / 0.19 |
| Middle | -0.72 / 1.08 / 1.2 / 0.33 | -0.67 / 1.14 / 1.7 / 0.38 | -0.80 / 1.17 / 1.8 / 0.26 |
| Fourth | -0.68 / 1.05 / 1.5 / 0.37 | -0.57 / 1.18 / 2.3 / 0.51 | -0.74 / 1.21 / 2.1 / 0.31 |
| Richest | -0.42 / 1.17 / 3.4 / 0.78 | -0.34 / 1.38 / 5.4 / 0.97 | -0.57 / 1.23 / 2.6 / 0.51 |

**S4 Table. Prevalence under alternative definitions, % (95% CI), and inequality in overweight without stunting.**

Overweight without stunting is WHZ above +2 with height-for-age at or above -2, among children with both valid scores.

| Group | 2012-13 | 2019 | 2025 |
| --- | --- | --- | --- |
| **BMI-for-age above +2** |  |  |  |
| Total | 1.7 (1.5 to 2.0) | 3.0 (2.7 to 3.3) | 2.1 (1.8 to 2.3) |
| Urban | 2.3 (1.6 to 3.2) | 5.9 (4.9 to 7.0) | 2.3 (1.8 to 2.8) |
| Rural | 1.6 (1.4 to 1.9) | 2.2 (2.0 to 2.5) | 2.0 (1.7 to 2.3) |
| Poorest | 1.1 (0.8 to 1.5) | 2.0 (1.6 to 2.4) | 0.9 (0.6 to 1.3) |
| Second | 1.1 (0.8 to 1.5) | 1.7 (1.3 to 2.2) | 1.7 (1.3 to 2.2) |
| Middle | 1.6 (1.1 to 2.2) | 2.0 (1.6 to 2.6) | 1.6 (1.3 to 2.2) |
| Fourth | 1.9 (1.4 to 2.5) | 3.0 (2.4 to 3.7) | 2.6 (2.1 to 3.3) |
| Richest | 3.3 (2.5 to 4.4) | 6.4 (5.4 to 7.5) | 3.4 (2.8 to 4.2) |
| **WHZ above +1** |  |  |  |
| Total | 5.7 (5.2 to 6.2) | 8.2 (7.8 to 8.7) | 6.0 (5.6 to 6.4) |
| Urban | 8.1 (6.7 to 9.8) | 13.4 (12.1 to 14.8) | 6.5 (5.8 to 7.3) |
| Rural | 5.0 (4.6 to 5.5) | 6.9 (6.5 to 7.4) | 5.8 (5.4 to 6.3) |
| Poorest | 4.3 (3.6 to 5.0) | 5.3 (4.7 to 6.0) | 3.7 (3.1 to 4.3) |
| Second | 4.1 (3.5 to 4.9) | 6.0 (5.2 to 6.8) | 4.3 (3.6 to 5.0) |
| Middle | 5.1 (4.2 to 6.0) | 6.7 (5.9 to 7.7) | 5.6 (4.9 to 6.5) |
| Fourth | 5.4 (4.5 to 6.4) | 8.8 (7.8 to 9.8) | 6.9 (6.1 to 7.9) |
| Richest | 10.2 (8.6 to 11.9) | 14.6 (13.2 to 16.0) | 9.6 (8.5 to 10.8) |
| **WHZ above +2, age 6-59 months** |  |  |  |
| Total | 1.3 (1.1 to 1.6) | 2.2 (2.0 to 2.5) | 1.6 (1.4 to 1.8) |
| Urban | 2.2 (1.5 to 3.1) | 4.8 (3.9 to 5.9) | 1.7 (1.3 to 2.2) |
| Rural | 1.1 (0.9 to 1.4) | 1.6 (1.3 to 1.8) | 1.6 (1.4 to 1.9) |
| Poorest | 0.6 (0.4 to 0.9) | 1.3 (1.0 to 1.8) | 0.8 (0.5 to 1.2) |
| Second | 0.9 (0.6 to 1.3) | 1.1 (0.8 to 1.6) | 1.1 (0.8 to 1.6) |
| Middle | 1.0 (0.6 to 1.5) | 1.4 (1.1 to 2.0) | 1.5 (1.1 to 2.0) |
| Fourth | 1.5 (1.1 to 2.0) | 2.0 (1.5 to 2.6) | 2.1 (1.7 to 2.7) |
| Richest | 3.0 (2.2 to 4.1) | 5.4 (4.4 to 6.5) | 2.5 (2.0 to 3.3) |
| **WHZ above +2, correct measurement position** |  |  |  |
| Total | 1.6 (1.4 to 1.9) | 2.4 (2.2 to 2.7) | 1.8 (1.5 to 2.0) |
| Urban | 2.8 (2.0 to 3.8) | 4.8 (4.0 to 5.9) | 1.8 (1.4 to 2.3) |
| Rural | 1.3 (1.1 to 1.5) | 1.8 (1.6 to 2.0) | 1.8 (1.5 to 2.0) |
| Poorest | 0.9 (0.7 to 1.2) | 1.5 (1.2 to 1.9) | 0.9 (0.6 to 1.3) |
| Second | 1.2 (0.9 to 1.6) | 1.3 (1.0 to 1.7) | 1.4 (1.0 to 1.9) |
| Middle | 1.2 (0.8 to 1.8) | 1.7 (1.3 to 2.2) | 1.8 (1.4 to 2.4) |
| Fourth | 1.5 (1.1 to 2.1) | 2.3 (1.8 to 2.9) | 2.2 (1.7 to 2.8) |
| Richest | 3.4 (2.6 to 4.6) | 5.4 (4.5 to 6.4) | 2.6 (2.0 to 3.2) |
| **WHZ above +2 and not stunted** |  |  |  |
| Total | 0.7 (0.5 to 0.9) | 1.1 (1.0 to 1.3) | 1.0 (0.8 to 1.2) |
| Urban | 1.3 (0.8 to 2.1) | 2.3 (1.8 to 2.9) | 1.1 (0.8 to 1.5) |
| Rural | 0.5 (0.4 to 0.7) | 0.8 (0.7 to 1.0) | 0.9 (0.8 to 1.1) |
| Poorest | 0.2 (0.1 to 0.4) | 0.5 (0.3 to 0.7) | 0.3 (0.2 to 0.6) |
| Second | 0.2 (0.1 to 0.3) | 0.4 (0.3 to 0.7) | 0.6 (0.4 to 0.9) |
| Middle | 0.4 (0.2 to 0.7) | 0.9 (0.6 to 1.3) | 0.8 (0.6 to 1.1) |
| Fourth | 0.6 (0.4 to 1.0) | 1.1 (0.8 to 1.6) | 1.3 (0.9 to 1.8) |
| Richest | 2.1 (1.4 to 3.0) | 2.9 (2.3 to 3.6) | 1.9 (1.4 to 2.5) |

| Overweight without stunting | 2012-13 | 2019 | 2025 |
| --- | --- | --- | --- |
| RII, crude | 45.89 (13.24 to 159.13) | 16.74 (8.63 to 32.48) | 9.61 (5.00 to 18.48) |
| RII, adjusted for residence | 41.33 (11.80 to 144.82) | 12.63 (6.37 to 25.06) | 12.02 (6.10 to 23.66) |
| Urban PR, adjusted for wealth rank | 1.12 (0.65 to 1.92) | 1.36 (0.98 to 1.89) | 0.71 (0.49 to 1.03) |
| Urban PR, crude | 2.65 (1.55 to 4.52) | 2.69 (1.97 to 3.67) | 1.19 (0.82 to 1.72) |

**S5 Table. Crude and adjusted prevalence ratios (95% CI) for overweight, 2019 and 2025.**

Modified Poisson models with survey design. Adjusted models include all variables shown. The area type model replaces residence with city corporation, other urban and rural.

| Variable | 2019 crude PR | 2019 adjusted PR | 2025 crude PR | 2025 adjusted PR |
| --- | --- | --- | --- | --- |
| **Age (months)** |  |  |  |  |
| 0-5 | 1 (reference) | 1 (reference) | 1 (reference) | 1 (reference) |
| 6-11 | 0.77 (0.54 to 1.10) | 0.77 (0.54 to 1.09) | 1.02 (0.70 to 1.47) | 1.03 (0.72 to 1.49) |
| 12-23 | 0.65 (0.47 to 0.90) | 0.67 (0.48 to 0.93) | 0.58 (0.40 to 0.86) | 0.60 (0.41 to 0.89) |
| 24-35 | 0.57 (0.40 to 0.80) | 0.57 (0.41 to 0.81) | 0.42 (0.28 to 0.62) | 0.43 (0.29 to 0.64) |
| 36-47 | 0.47 (0.33 to 0.66) | 0.49 (0.35 to 0.69) | 0.50 (0.33 to 0.75) | 0.52 (0.35 to 0.79) |
| 48-59 | 0.34 (0.23 to 0.49) | 0.35 (0.24 to 0.52) | 0.42 (0.28 to 0.63) | 0.46 (0.30 to 0.69) |
| **Sex** |  |  |  |  |
| Male | 1 (reference) | 1 (reference) | 1 (reference) | 1 (reference) |
| Female | 0.87 (0.70 to 1.08) | 0.88 (0.72 to 1.09) | 0.89 (0.71 to 1.13) | 0.86 (0.68 to 1.08) |
| **Mother’s education** |  |  |  |  |
| None or pre-primary | 1 (reference) | 1 (reference) | 1 (reference) | 1 (reference) |
| Primary | 1.23 (0.80 to 1.90) | 1.07 (0.69 to 1.66) | 0.71 (0.38 to 1.33) | 0.63 (0.34 to 1.17) |
| Secondary | 1.47 (1.01 to 2.15) | 1.12 (0.76 to 1.66) | 0.70 (0.41 to 1.20) | 0.56 (0.33 to 0.96) |
| Higher secondary or above | 2.73 (1.79 to 4.15) | 1.43 (0.91 to 2.23) | 1.49 (0.85 to 2.62) | 1.06 (0.58 to 1.90) |
| **Wealth quintile** |  |  |  |  |
| Poorest | 1 (reference) | 1 (reference) | 1 (reference) | 1 (reference) |
| Second | 0.83 (0.57 to 1.20) | 0.76 (0.53 to 1.11) | 1.51 (0.93 to 2.44) | 1.46 (0.91 to 2.34) |
| Middle | 1.10 (0.76 to 1.60) | 0.99 (0.68 to 1.44) | 1.97 (1.26 to 3.07) | 1.71 (1.10 to 2.65) |
| Fourth | 1.47 (1.03 to 2.10) | 1.15 (0.79 to 1.66) | 2.40 (1.57 to 3.66) | 1.91 (1.23 to 2.96) |
| Richest | 3.51 (2.57 to 4.78) | 2.06 (1.47 to 2.89) | 2.89 (1.88 to 4.47) | 2.08 (1.29 to 3.35) |
| **Residence** |  |  |  |  |
| Rural | 1 (reference) | 1 (reference) | 1 (reference) | 1 (reference) |
| Urban | 2.69 (2.13 to 3.40) | 1.52 (1.19 to 1.93) | 0.99 (0.75 to 1.31) | 0.65 (0.48 to 0.87) |
| **Division** |  |  |  |  |
| Barishal | 1 (reference) | 1 (reference) | 1 (reference) | 1 (reference) |
| Chattogram | 0.94 (0.59 to 1.49) | 0.74 (0.46 to 1.20) | 0.89 (0.53 to 1.51) | 0.80 (0.47 to 1.37) |
| Dhaka | 2.43 (1.58 to 3.74) | 1.61 (1.04 to 2.47) | 2.53 (1.61 to 3.98) | 2.29 (1.43 to 3.67) |
| Khulna | 0.67 (0.38 to 1.20) | 0.60 (0.34 to 1.06) | 0.80 (0.45 to 1.40) | 0.72 (0.41 to 1.27) |
| Mymensingh | 0.83 (0.45 to 1.55) | 0.87 (0.47 to 1.60) | 0.99 (0.46 to 2.12) | 1.04 (0.49 to 2.22) |
| Rajshahi | 0.92 (0.55 to 1.53) | 0.86 (0.51 to 1.44) | 0.92 (0.52 to 1.66) | 0.85 (0.48 to 1.53) |
| Rangpur | 1.23 (0.75 to 2.02) | 1.29 (0.78 to 2.14) | 0.64 (0.35 to 1.16) | 0.60 (0.33 to 1.09) |
| Sylhet | 0.52 (0.27 to 0.98) | 0.45 (0.23 to 0.87) | 1.20 (0.62 to 2.33) | 1.07 (0.56 to 2.05) |
| **Household members** |  |  |  |  |
| 1-4 | 1 (reference) | 1 (reference) | 1 (reference) | 1 (reference) |
| 5-6 | 0.83 (0.66 to 1.04) | 0.92 (0.73 to 1.15) | 0.94 (0.72 to 1.22) | 0.97 (0.74 to 1.27) |
| 7 or more | 0.95 (0.73 to 1.24) | 0.98 (0.75 to 1.28) | 1.02 (0.73 to 1.42) | 0.95 (0.69 to 1.32) |
| **Religion of household head** |  |  |  |  |
| Other religion | 1 (reference) | 1 (reference) | 1 (reference) | 1 (reference) |
| Islam | 1.55 (1.03 to 2.33) | 1.31 (0.87 to 1.98) | 0.70 (0.44 to 1.11) | 0.63 (0.40 to 0.99) |
| **Area type, 2025 (adjusted, rural reference)** |  |  |  |  |
| City corporation |  |  |  | 0.46 (0.27 to 0.78) |
| Other urban |  |  |  | 0.72 (0.53 to 0.98) |

**S6 Table. Urban Dhaka district, other urban and rural areas, and wealth standardisation of urban prevalence.**

Urban Dhaka district covers urban clusters of Dhaka district, including Dhaka North and Dhaka South City Corporations in 2025. Population share is the weighted share of measured children.

| Wave | Group | Children (clusters) | Overweight cases | Overweight, % (95% CI) | Mean WHZ | SD of WHZ | Population share, % | Cases in top five clusters, % |
| --- | --- | --- | --- | --- | --- | --- | --- | --- |
| 2012-13 | Urban Dhaka district | 156 (31) | 5 | 3.6 (1.5 to 8.4) | -0.40 | 1.15 | 4.5 | 100.0 |
| 2012-13 | Other urban | 2,970 (422) | 70 | 2.5 (1.8 to 3.4) | -0.66 | 1.15 | 15.9 | 15.7 |
| 2012-13 | Rural | 16,472 (2,298) | 228 | 1.3 (1.1 to 1.5) | -0.72 | 1.08 | 79.6 | 7.5 |
| 2019 | Urban Dhaka district | 526 (96) | 67 | 12.1 (9.1 to 15.9) | 0.01 | 1.66 | 5.0 | 23.9 |
| 2019 | Other urban | 3,517 (536) | 80 | 2.6 (2.0 to 3.3) | -0.55 | 1.20 | 15.9 | 15.0 |
| 2019 | Rural | 18,020 (2,582) | 309 | 1.8 (1.6 to 2.1) | -0.69 | 1.17 | 79.2 | 6.8 |
| 2025 | Urban Dhaka district | 629 (103) | 16 | 2.3 (1.4 to 3.9) | -0.62 | 1.21 | 4.9 | 43.8 |
| 2025 | Other urban | 5,290 (755) | 83 | 1.6 (1.2 to 2.1) | -0.77 | 1.16 | 23.8 | 13.3 |
| 2025 | Rural | 16,392 (2,286) | 269 | 1.7 (1.5 to 2.0) | -0.81 | 1.18 | 71.3 | 5.6 |

Urban overweight (%) standardised to the urban wealth-quintile mix of each wave (columns)

| Quintile-specific urban rates from | 2012-13 | 2019 | 2025 |
| --- | --- | --- | --- |
| Rates 2012-13 | 2.74 | 2.92 | 2.49 |
| Rates 2019 | 4.50 | 4.84 | 4.06 |
| Rates 2025 | 1.83 | 1.92 | 1.73 |

| Wave | Quintile | Urban overweight, % | Share of urban children, % | Rural overweight, % | Share of rural children, % |
| --- | --- | --- | --- | --- | --- |
| 2012-13 | Poorest | 0.3 | 10.1 | 0.9 | 28.1 |
| 2012-13 | Second | 2.3 | 9.0 | 1.1 | 23.4 |
| 2012-13 | Middle | 0.7 | 11.8 | 1.3 | 20.5 |
| 2012-13 | Fourth | 1.7 | 21.6 | 1.4 | 17.1 |
| 2012-13 | Richest | 4.3 | 47.6 | 2.4 | 10.9 |
| 2019 | Poorest | 1.1 | 7.0 | 1.6 | 25.6 |
| 2019 | Second | 0.6 | 6.8 | 1.3 | 23.2 |
| 2019 | Middle | 2.6 | 11.0 | 1.6 | 20.8 |
| 2019 | Fourth | 2.8 | 22.4 | 2.1 | 18.8 |
| 2019 | Richest | 7.2 | 52.8 | 3.2 | 11.5 |
| 2025 | Poorest | 0.6 | 9.7 | 0.9 | 24.2 |
| 2025 | Second | 1.0 | 10.7 | 1.4 | 23.6 |
| 2025 | Middle | 1.5 | 14.8 | 1.8 | 22.0 |
| 2025 | Fourth | 1.5 | 26.5 | 2.5 | 18.6 |
| 2025 | Richest | 2.5 | 38.3 | 2.7 | 11.7 |

**S7 Table. Concurrent overweight and stunting.**

Cell: % of all children overweight and stunted; % of overweight children who are stunted (95% CI) [overweight children with valid HAZ]

| Group | 2012-13 | 2019 | 2025 |
| --- | --- | --- | --- |
| Total | 0.74; 52.7 (43.9 to 61.4) [n = 259] | 1.13; 49.7 (44.2 to 55.3) [n = 422] | 0.69; 41.4 (35.3 to 47.7) [n = 348] |
| Poorest | 0.53; 70.6 (53.8 to 83.2) [n = 55] | 0.83; 64.1 (50.9 to 75.6) [n = 69] | 0.45; 57.9 (39.2 to 74.6) [n = 42] |
| Second | 0.72; 80.6 (66.1 to 89.8) [n = 43] | 0.79; 65.4 (50.9 to 77.5) [n = 55] | 0.72; 54.8 (40.7 to 68.2) [n = 59] |
| Middle | 0.58; 58.4 (39.4 to 75.2) [n = 38] | 0.74; 45.2 (31.9 to 59.3) [n = 61] | 0.85; 51.5 (38.4 to 64.4) [n = 74] |
| Fourth | 0.63; 49.4 (31.9 to 67.0) [n = 44] | 1.09; 49.6 (37.1 to 62.1) [n = 84] | 0.74; 36.3 (25.2 to 49.1) [n = 86] |
| Richest | 1.32; 39.0 (24.3 to 56.1) [n = 79] | 2.19; 43.4 (34.3 to 53.0) [n = 153] | 0.68; 26.7 (18.0 to 37.6) [n = 87] |
| Rural | 0.58; 53.9 (44.6 to 62.9) [n = 188] | 0.82; 49.3 (42.7 to 55.9) [n = 283] | 0.73; 44.0 (36.6 to 51.6) [n = 252] |
| Urban | 1.37; 51.0 (33.9 to 67.8) [n = 71] | 2.30; 50.3 (40.8 to 59.8) [n = 139] | 0.59; 35.0 (25.0 to 46.5) [n = 96] |

**S8 Table. Sensitivity analysis excluding urban Dhaka district, and overweight cases by measurer.**

In 2019, one measurer measured 508 of the 526 children in urban Dhaka district and recorded all 67 overweight cases. Measurer codes are those in the public data files.

| Group or measure | 2012-13 | 2019 | 2025 |
| --- | --- | --- | --- |
| Total | 1.5 (1.3 to 1.7) | 1.9 (1.7 to 2.2) | 1.7 (1.5 to 1.9) |
| Rural | 1.3 (1.1 to 1.5) | 1.8 (1.6 to 2.1) | 1.7 (1.5 to 2.0) |
| Urban | 2.5 (1.8 to 3.4) | 2.6 (2.0 to 3.3) | 1.6 (1.2 to 2.1) |
| Poorest | 0.9 (0.7 to 1.2) | 1.5 (1.2 to 2.0) | 0.9 (0.6 to 1.3) |
| Second | 1.2 (0.9 to 1.7) | 1.3 (0.9 to 1.7) | 1.3 (1.0 to 1.8) |
| Middle | 1.2 (0.9 to 1.8) | 1.7 (1.3 to 2.2) | 1.8 (1.4 to 2.3) |
| Fourth | 1.5 (1.1 to 2.1) | 2.0 (1.6 to 2.6) | 2.3 (1.8 to 2.9) |
| Richest | 3.1 (2.4 to 4.2) | 3.4 (2.8 to 4.2) | 2.4 (1.9 to 3.1) |
| RII crude | 4.87 (2.68 to 8.86) | 3.11 (1.96 to 4.94) | 3.58 (2.29 to 5.59) |
| RII mutually adjusted | 4.03 (2.25 to 7.24) | 2.96 (1.82 to 4.82) | 4.16 (2.62 to 6.62) |
| Urban PR mutually adjusted | 1.43 (1.01 to 2.03) | 1.10 (0.82 to 1.47) | 0.71 (0.52 to 0.97) |
| Urban PR crude | 1.93 (1.36 to 2.73) | 1.42 (1.08 to 1.87) | 0.92 (0.68 to 1.25) |

Change in overweight excluding urban Dhaka district, percentage points: 2019 vs 2012-13 0.43 (0.12 to 0.75); 2025 vs 2019 -0.21 (-0.53 to 0.10) (p = 0.180).
Wealth rank by wave interaction p = 0.507; urban by wave interaction p = 0.006.

| Wave | Area | Measurers | Children | Overweight cases | Cases recorded by the two busiest measurers, % |
| --- | --- | --- | --- | --- | --- |
| 2019 | Urban Dhaka district | 4 | 526 | 67 | 100.0 |
| 2019 | Elsewhere | 96 | 21,537 | 389 | 23.1 |
| 2025 | Urban Dhaka district | 3 | 629 | 16 | 93.8 |
| 2025 | Elsewhere | 41 | 21,682 | 352 | 18.8 |

**S9 Table. Participant flow, and children with and without a valid WHZ.**

Weighted percentages among children with a completed child questionnaire.

| Step | 2012-13 | 2019 | 2025 |
| --- | --- | --- | --- |
| Children under five listed in sampled households | 23,402 | 24,686 | 24,680 |
| Child questionnaire completed | 20,903 | 23,099 | 23,357 |
| Weight and height recorded | 19,762 | 22,266 | 22,491 |
| Excluded: implausible WHZ (flagged) | 164 | 203 | 180 |
| Analytic sample (valid WHZ) | 19,598 | 22,063 | 22,311 |

Weighted % of children with a valid WHZ / without a valid WHZ, among those with a completed child questionnaire. Children without a valid WHZ: 1,305 (2012-13); 1,036 (2019); 1,046 (2025).

| Group | 2012-13 | 2019 | 2025 |
| --- | --- | --- | --- |
| **Age (months)** |  |  |  |
| 0-5 | 9.3 / 12.3 | 10.1 / 16.6 | 9.4 / 15.1 |
| 6-11 | 9.8 / 5.7 | 9.8 / 4.2 | 9.8 / 6.5 |
| 12-23 | 20.0 / 13.3 | 19.6 / 12.1 | 21.0 / 19.1 |
| 24-35 | 19.6 / 26.7 | 19.6 / 26.1 | 21.0 / 21.4 |
| 36-47 | 20.6 / 22.8 | 20.8 / 21.8 | 19.7 / 19.1 |
| 48-59 | 20.7 / 19.2 | 20.1 / 19.2 | 19.1 / 18.8 |
| **Sex** |  |  |  |
| Male | 51.2 / 51.2 | 52.0 / 51.4 | 51.8 / 52.8 |
| Female | 48.8 / 48.8 | 48.0 / 48.6 | 48.2 / 47.2 |
| **Residence** |  |  |  |
| Rural | 79.6 / 79.8 | 79.2 / 70.9 | 71.3 / 70.7 |
| Urban | 20.4 / 20.2 | 20.8 / 29.1 | 28.7 / 29.3 |
| **Wealth quintile** |  |  |  |
| Poorest | 24.5 / 23.9 | 21.8 / 22.8 | 20.0 / 19.6 |
| Second | 20.4 / 21.4 | 19.7 / 17.3 | 19.9 / 16.7 |
| Middle | 18.7 / 16.9 | 18.8 / 14.9 | 19.9 / 21.6 |
| Fourth | 18.0 / 16.9 | 19.6 / 18.7 | 20.9 / 20.0 |
| Richest | 18.4 / 20.9 | 20.1 / 26.3 | 19.3 / 22.1 |
| **Mother’s education** |  |  |  |
| None or pre-primary | 22.3 / 24.7 | 11.1 / 12.5 | 6.0 / 5.5 |
| Primary | 29.8 / 28.2 | 23.7 / 24.2 | 17.9 / 19.1 |
| Secondary or higher | 47.9 / 47.1 | 65.2 / 63.3 | 76.2 / 75.3 |

**S10 Table. Main estimates with response weights for missing or implausible measurement, % or ratio (95% CI).**

Response weights multiply the survey weight by the inverse of the modelled probability of a valid WHZ, given age group, sex, residence, wealth quintile, division and mother’s education.

| Estimate | 2012-13 | 2019 | 2025 |
| --- | --- | --- | --- |
| Overweight, %: Total | 1.6 (1.4 to 1.9) | 2.4 (2.2 to 2.7) | 1.7 (1.5 to 2.0) |
| Overweight, %: Urban | 2.8 (2.0 to 3.8) | 4.8 (4.0 to 5.8) | 1.7 (1.4 to 2.2) |
| Overweight, %: Rural | 1.3 (1.1 to 1.6) | 1.8 (1.6 to 2.1) | 1.7 (1.5 to 2.0) |
| Overweight, %: Poorest | 0.9 (0.7 to 1.3) | 1.5 (1.2 to 2.0) | 0.9 (0.6 to 1.3) |
| Overweight, %: Richest | 3.4 (2.6 to 4.6) | 5.4 (4.5 to 6.4) | 2.6 (2.0 to 3.2) |
| RII, fully adjusted | 3.59 (1.90 to 6.77) | 3.52 (2.19 to 5.66) | 3.34 (2.01 to 5.53) |
| Urban PR, adjusted for wealth rank | 1.44 (1.01 to 2.06) | 1.76 (1.40 to 2.23) | 0.72 (0.54 to 0.96) |
